# Safety-Relevant Biomedical Machine Learning Should Adopt Stability-First Reporting

**DOI:** 10.64898/2026.09.30.26364447

**Authors:** Zayn Andre Zainal, Omid Kavehei, Isabelle Aguilar, Luis Fernando Herbozo Contreras, Zhaojing Huang

## Abstract

Biomedical Machine Learning (ML) is increasingly evaluated through Independent and Identically Distributed (IID) benchmarks, even when deployment involves irregular observation, missingness, distribution shift, and high failure costs. This position paper argues for a stability-oriented reporting standard for safety-relevant biomedical ML. The central requirement is not a specific model class or optimizer, but auditable evidence: performance should be reported under clinically plausible perturbations such as thinning, timestamp jitter, bursty missingness, channel dropout, and Signal-to-Noise Ratio (SNR) shift. When authors invoke training-stability diagnostics, those diagnostics should be hazard-linked: their assumptions, operational meaning, and relationship to stress-test degradation should be stated explicitly. We use controlled mechanism probes to illustrate two hazards that IID scores can hide: observation-process dependence under irregular sampling, and perturbation sensitivity associated with high-curvature training regimes such as Edge of Stability (EoS). We do not propose Neural ODEs, continuous-time models, Sharpness-Aware Minimization (SAM), or EoS metrics as default prescriptions. Instead, we propose a minimum reporting standard that makes robustness claims testable and supports escalation to more complex modelling or optimization only when stress tests justify the cost.

## 1 Introduction

Biomedical Artificial Intelligence (AI) increasingly informs decisions in settings where failure is unacceptable. Imaging reconstruction, acute event prediction, and closed-loop intervention are key areas in the clinical industry (to name a few) where AI inference is becoming more mainstream [1, 2]. Yet, the dominant pipeline still optimizes Independent and Identically Distributed (IID) predictive performance under discrete-time supervision [3, 4]. *Finlayson et al*. [5] assert that clinical AI failures often come from data set changes rather than capacity modeling alone, motivating stress-tested evaluation and regime shift monitoring to ensure valid deployment. Stable behavior under irregular observation, distribution shift, and physiological non-stationarity are rarely targeted [5–7]. We argue that this gap is structural. Many failures have been linked to mismatches in temporal assumptions and curvature-mediated training dynamics, not only to limited data or insufficient capacity [8, 9].

### Position: Safety-relevant biomedical Machine Learning (ML) should be evaluated through a stability-oriented reporting standard, not a mandatory modelling recipe

The required evidence is (i) schedule-perturbation stress testing under clinically plausible observation shifts and (ii) hazard-linked training diagnostics when such diagnostics are used to support robustness claims. By *hazard-linked training diagnostic*, we mean a training-time quantity whose relevance is evaluated against a predefined deployment-relevant stress test, rather than treated as a safety certificate by itself.

We use seizure forecasting as a motivating example because it is *transition-driven*: clinically relevant risk concentrates around an impending state change rather than stationary pattern recognition. Although seizure onset is often framed as a dynamical transition [10], many models remain fragile under clinically realistic shifts such as sampling jitter, channel dropout, or SNR degradation, and this brittleness can be exacerbated when discrete-time assumptions interact with Edge-of-Stability training (for example, sustained *ρ*(*t*) *>* 1) [11, 12]. More broadly, shortcut learning and weak external validity are recurrent in medical AI [5, 9, 13, 14], so we present our results not as clinical validation but rather as *mechanism probes* to motivate the need for a minimum-risk reporting standard. By *mechanism probe*, we mean a controlled experiment designed to isolate one failure mechanism, not an end-to-end clinical validation study.

This position paper advances the following reporting agenda for the NeurIPS community:

- **Position 1 (Irregular time):** Report performance under thinning, jitter, and bursty missingness; use continuous-time bias or principled discretization when needed. Here, *thinning* means systematically removing a fraction of observations, *jitter* means perturbing observation timestamps while keeping measured values fixed, and *bursty missingness* means removing contiguous time intervals rather than independent missing-at-random points.
- **Position 2 (Optimization stability):** When training-instability claims are made, report the diagnostic used, its assumptions, and its link to stress-test degradation. One possible diagnostic is the normalized curvature ratio *ρ*(*t*) = *η*(*t*)*λ*_max_(*t*)*/*2, where *η*(*t*) is the learning rate and *λ*_max_(*t*) is a top Hessian-eigenvalue proxy, typically estimated on a fixed *probe batch* that is not used for optimization. We use *T*_EoS_ to denote the empirical fraction of training steps or epochs for which *ρ*(*t*) *>* 1.
- **Position 3 (Regime changes):** For transition-driven tasks, report lead time at a fixed false alarm rate using latent monitoring signals. Lead time is the interval between the first valid alarm and the true transition point, where a valid alarm is one that occurs inside a predefined pre-event window. False alarm rate is the rate of alarms outside that window, normalized per unit time or monitoring episode.

These positions align closely with *Grote et al*. [14], explaining that foundation models require rethinking the reliability evaluation process. They specifically call for new reliability checks for healthcare use, including evaluations that are more aware of shift, context, and failure mode.

## 2 Gap A: The Modeling Hazard of Irregular Clinical Data

Biological systems evolve in continuous time, yet our empirical observations are fundamentally constrained by a discrete sampling process. Biological processes ranging from exponential drug clearance to the propagation of electrical membrane potentials are fundamentally governed by differential equations. For example, a fundamental law of electrophysiology is

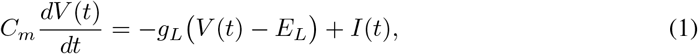

where *V* (*t*) is the membrane potential, *C*_*m*_ is the membrane capacitance, *g*_*L*_ and *E*_*L*_ are the leak conductance and the reversal potential, and *I*(*t*) is the net input current. Nevertheless, in practice,clinical measurements are recorded at finite and often variable rates, with gaps in data and irregular timing that can depend on workflow and patient state. Similarly, canonical deep learning architectures are typically formulated in discrete time [15, 16]. Although Long Short-Term Memory (LSTM) networks can achieve high sensitivity in applications such as seizure prediction by capturing temporal dependencies [17, 18], fixed-step assumptions can become a structural weakness when the underlying physiology is continuous, the sampling schedule varies, and this dependence is not audited.

**Figure 1:**
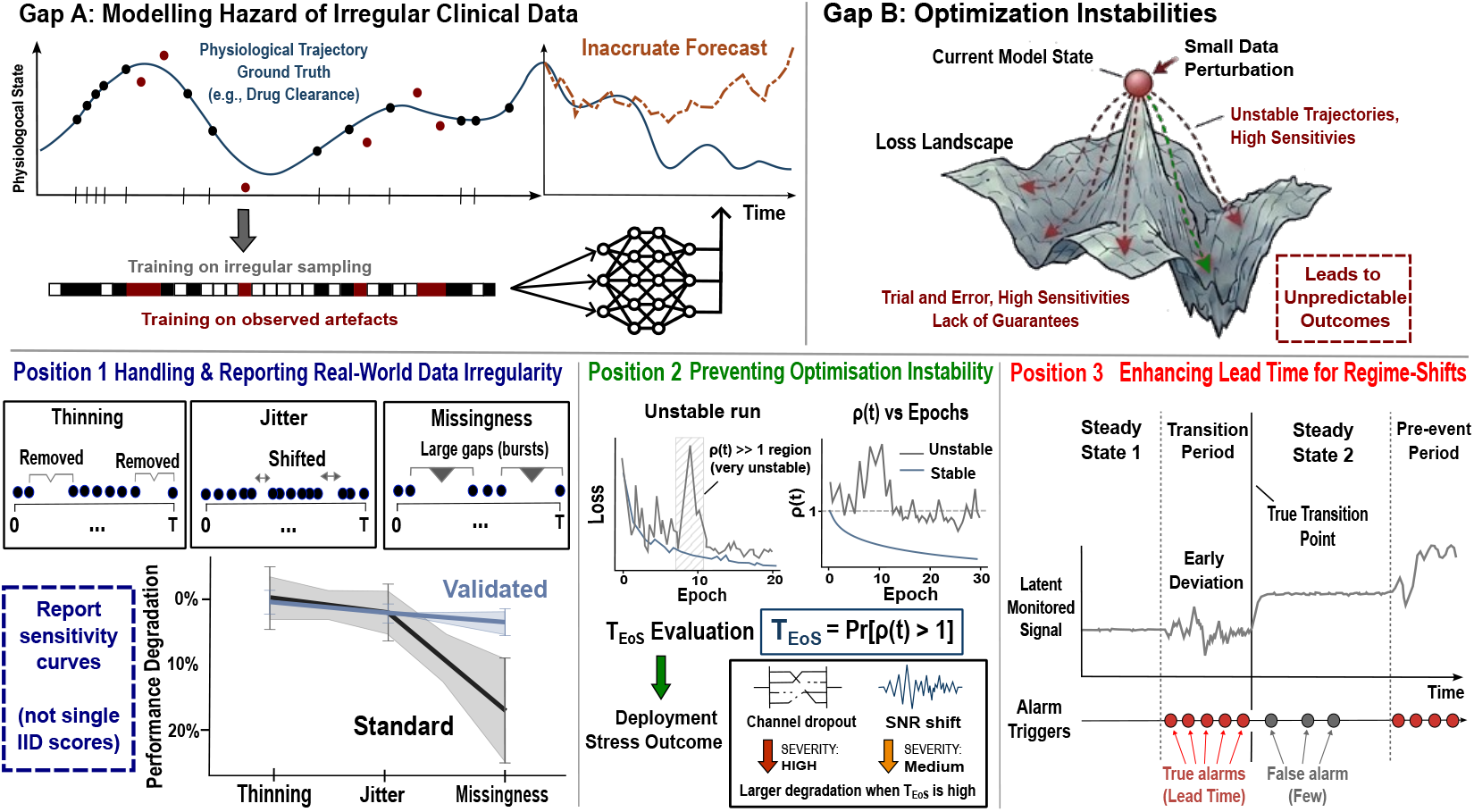
Stability-first biomedical ML targets two hazards missed by IID, discrete-time defaults. *(Gap A)* Irregular sampling can conflate physiology with the observation process, so we report degradation slopes under schedule perturbations (not single-point IID scores). *(Gap B)* Training can remain near or beyond a local stability boundary (*ρ*(*t*) = *ηλ*_max_(*t*)*/*2 *>* 1); when robustness or optimization-stability claims are made, reporting *ρ*(*t*) and *T*_EoS_ = Pr[*ρ*(*t*) *>* 1] can make those claims auditable. For transition-driven tasks, reports should also specify false-alarm rate and lead time. Elements of this figure were initially drafted using *Notion AI* and subsequently refined by the authors to ensure representational clarity.

This issue arises in several common biomedical settings. In EEG seizure forecasting, models trained on regular windows can fail when deployed under clinically realistic channel dropout, artifact rejection, variable electrode quality, or SNR degradation; the model may learn recording regularity rather than pre-ictal dynamics. In ICU deterioration prediction, measurement frequency often changes with patient acuity, so a fixed-step model can treat the observation schedule itself as a severity shortcut rather than learning physiology. In pharmacokinetic or physiological monitoring, sparse follow-up and interpolation can make synthetic imputation artifacts appear as disease dynamics, especially when samples are thinned, jittered, or missing in bursts. These settings motivate reporting degradation curves under thinning, jitter, bursty missingness, channel dropout, and SNR shift, rather than relying only on single IID scores.

### 2.1 When Fixed-Step Pipelines Hide Observation-Process Risk

The problem is not discrete-time modelling itself. Discrete-time methods are often appropriate when measurements are regular, horizons are short, and stress-test degradation is small. The risk arises when irregularly observed physiology is forced into fixed-step pipelines without auditing the observation process. Standard Recurrent Neural Network (RNN)-based sequence models often assume observations arrive on a regular grid. For example, in Electroencephalography (EEG), modern sampling rates range from 256 to 512 Hz [19, 20]. In clinical practice, however, measurements are frequently sparse, intermittent, and irregular [21]. Converting such data into fixed-step inputs typically requires interpolation, imputation, windowing, or masking choices [22, 23]. These choices are not neutral: they can introduce artifacts or allow the model to learn the observation schedule rather than the latent disease trajectory. This creates a non-obvious failure mode: apparently strong IID performance, but brittle behavior when sampling frequency, timing, or missingness mechanisms shift at deployment.

### 2.2 Continuous-Time Models as One Irregular-Time Tool

Continuous-time models are useful when the timing of observations is itself part of the modelling problem. A Neural ODE, for example, represents the hidden state *h*(*t*) as the solution to an initial value problem,

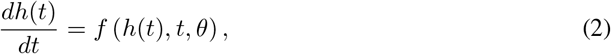

so predictions can be evaluated at arbitrary observation times rather than only on a fixed grid [19, 20]. This makes NODEs a natural tool for auditing irregular sampling, but it does not make them a default requirement. Discrete-time models with explicit time encodings, masking, learned interpolation, or principled continuous-to-discrete constructions may be preferable when they achieve comparable stress-test robustness at lower computational cost.

We use FitzHugh-Nagumo dynamics as a controlled *mechanism probe* for this issue (Fig. 3, Appendix A.2). The value of this setting is that the ground-truth trajectory is known and the observation schedule can be perturbed systematically. Under irregular resampling, the discrete sequence baselines in our experiment progressively accumulate phase error, whereas the NODE remains closer to the limit-cycle trajectory and keeps long-horizon error bounded. This result should be read as evidence that observation-schedule dependence can be audited, not as proof that NODEs dominate discrete-time models in clinical deployment.

Accordingly, the reporting requirement is model-agnostic: when a biomedical ML claim depends on temporal robustness, authors should report performance under schedule perturbations and compare against appropriate baselines under matched capacity and compute budgets. For long, high-rate signals, solver-based NODEs can be costly; State Space Models (SSMs; S4/Mamba-style) [24–26] or strong time-aware recurrent baselines may be more practical. These alternatives should be evaluated with the same irregular-time and shift-sensitivity tests. For reference, Table 4 summarizes the core dynamical-systems concepts used throughout the paper and their biomedical interpretations.

### 2.3 Regime Shifts and Early-Warning Evaluation

Many biomedical events of interest are not well modeled as stationary pattern-recognition problems but as *regime shifts* in an underlying dynamical system [10]. In this lens, clinically “normal” dynamics correspond to stable attractors (e.g., fixed points for homeostasis or limit cycles for rhythms), while acute events correspond to qualitative changes in phase-space geometry (bifurcations) [10, 27]. **This frame of reference motivates the evaluation of forecasting and early warning models by their behavior under perturbations and shifts, not only by IID classification metrics**.

A classical precursor for approaching critical transitions is *Critical Slowing Down* (CSD): near a bifurcation, perturbations decay more slowly, often increasing autocorrelation and variance [28]. We treat CSD as a working hypothesis, not a universally established biomarker in all modalities. Its real utility lies in the discipline it imposes: it translates a qualitative idea into a concrete monitoring criterion that can be tested and, if necessary, refuted. **Mechanism probe;** We simulate a supercritical Hopf bifurcation with slowly ramped *µ*(*t*) crossing *µ* = 0 under small process noise;see Appendix A.4. In this simulation, we observe rolling AR(1) and Var(*z*_*t*_) on *detrended residuals* rise as *µ*(*t*) *→* 0^*−*^, before sustained oscillations (Fig. 6). This is a falsifiable monitoring template, not a universal biomedical claim (Prediction 3 and Section 6).

## 3 Gap B: The Risk of Optimization Instability

Predictive precision is necessary but insufficient in high-stakes biomedical environments. A deployed model must remain stable under perturbations and distribution changes [29]. We treat Edge of Stability (EoS) behavior as a *conditional diagnostic*: it is informative only when a paper claims optimization stability, robustness under shift, or reliability under perturbation. Classical convex optimization analyses presume stable discrete-time descent in locally quadratic basins. By contrast, modern deep networks can exhibit *progressive sharpening*: curvature increases over training until the optimizer operates near or beyond a local quadratic stability boundary, while the training loss can still decrease [11, 30, 31]. This creates a metric ambiguity rather than a safety verdict: decreasing loss does not by itself establish stable local geometry, and curvature spikes do not by themselves prove deployment failure. The relevant question is whether high-curvature training behavior predicts a pre-specified degradation pattern under domain-relevant stress tests.

### Mechanism probe

We observe EoS-like behavior on PathMNIST histopathology data (Fig. 2). Although PathMNIST is not a deployment benchmark for clinical forecasting, it is useful for measurement clarity and reproducibility. The operational hypothesis is modality-agnostic but conditional: curvature-selected solutions may be more perturbation-sensitive under shift, and this should be tested rather than assumed. The sharpness proxy *λ*_max_, estimated as the largest Hessian eigenvalue on a fixed probe batch, exceeds the nominal stability threshold 2*/η* in our probe; a *probe batch* is a fixed mini-batch used only for curvature estimation, distinct from the training batches used for optimization. We therefore use this experiment to motivate diagnostic reporting, not to claim that EoS behavior is itself a clinical safety failure.IID
score, time-to-target, tuning effort, and degradation under clinically plausible perturbations.Table 5 summarises optimiser families as reporting-relevant design choices rather than a ranking.

**Figure 2:**
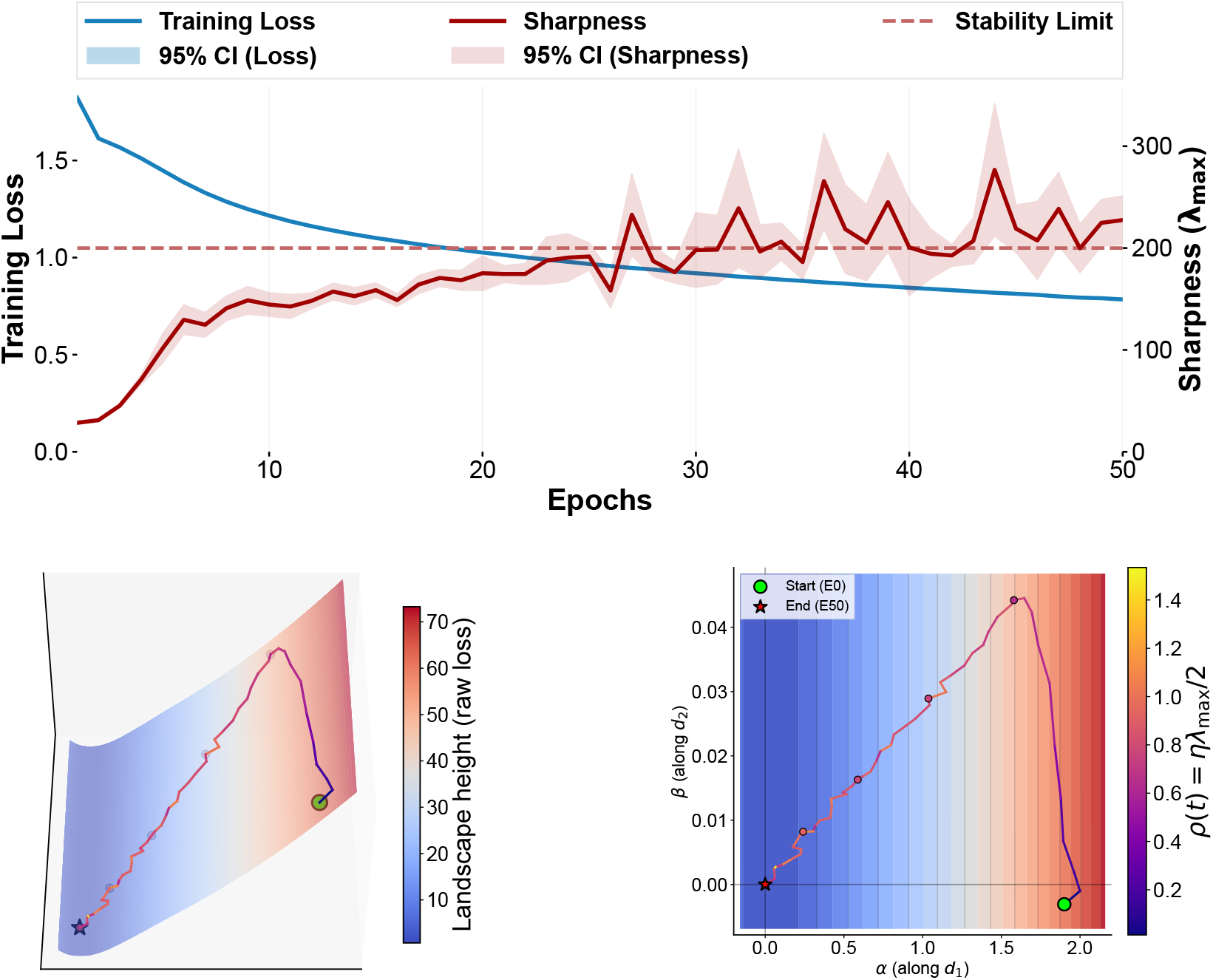
Edge of Stability dynamics in neural network optimization on PathMNIST.

**Figure 3:**
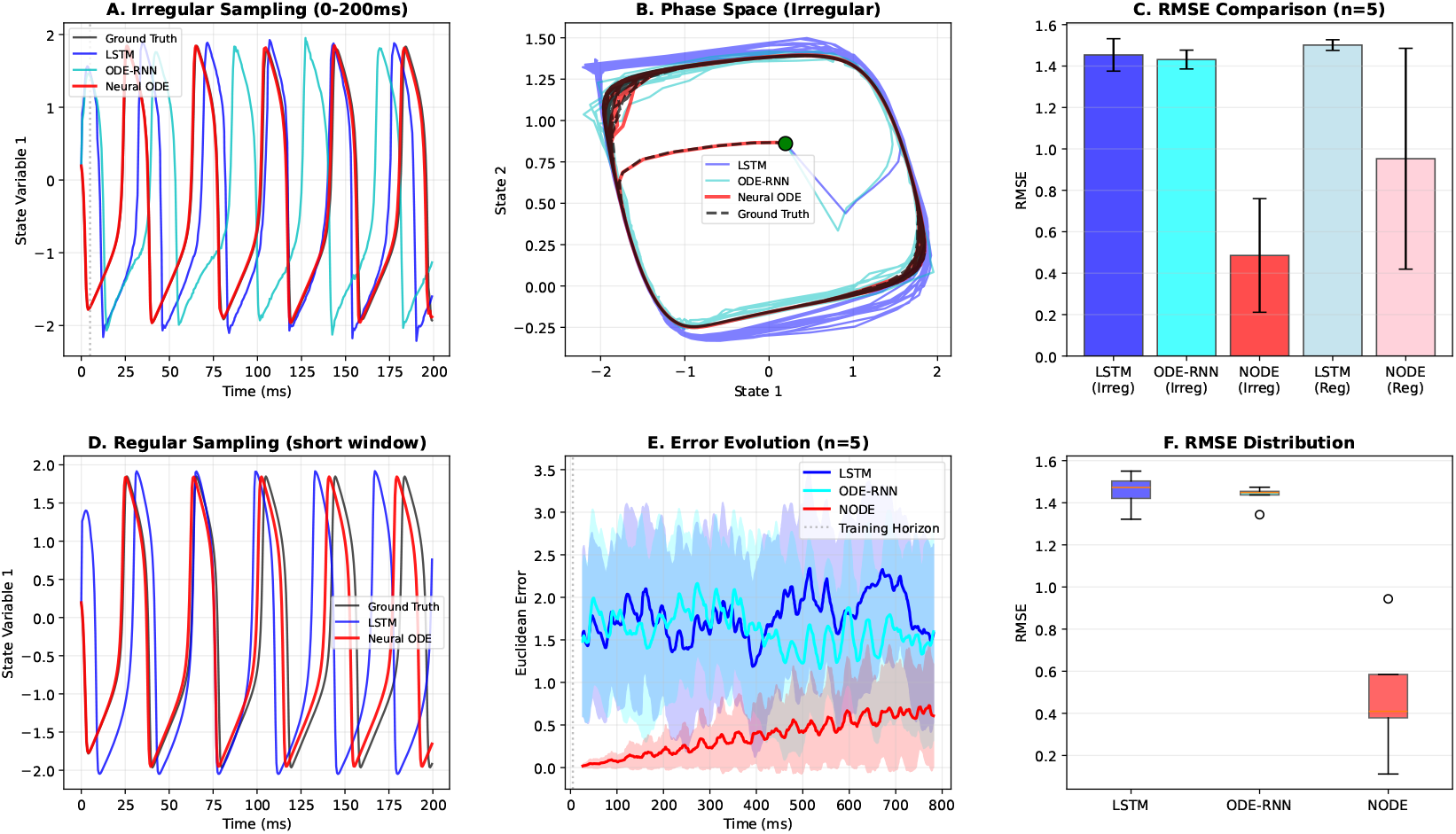
Neural ODEs reduce cumulative drift in this FitzHugh-Nagumo irregular-sampling probes. (**a**) Time-domain trajectories (0-200 ms) under irregular sampling (Δ*t* = 0.5 *±*0.3 ms training, 0.5 *±* 0.45 ms test). The time-aware LSTM (blue) and ODE-RNN (cyan) desynchronize due to error accumulation, whereas the Neural ODE (red) maintains phase-locked oscillations. (**b**) Phase-space trajectories show discrete models diverging from the limit cycle attractor, while the Neural ODE converges to and maintains stable periodic dynamics. (**c**) RMSE comparison. (**d**) Regular sampling control. (**e**) Temporal error evolution shows Neural ODE error remains bounded (*<* 1.0) throughout the 800 ms extrapolation. Shaded regions denote 1*±* s.d. across *n* = 5 independent random seeds (computed pointwise over time and smoothed with the same moving-average window as the mean). (**f**) RMSE distribution boxplots.

### Loss landscape topology should be treated as a conditional diagnostic

The classical stability condition follows from the linearization in a locally quadratic basin, producing *λ*_max_ *≤*2*/η*. The derivation and assumptions are given in Appendix D.1. Although deep network training is globally non-convex, the boundary remains diagnostically useful because it indicates when the update is unstable along the sharpest local direction. A mechanistic account of how EoS training can still reduce loss while violating the local bound is summarized in Appendix D.2.

Because deep-network objectives are non-convex, the quadratic-basin condition *λ*_max_ *≤* 2*/η* is not a convergence guarantee, nor does violating it prove deployment failure. We therefore use *ρ*(*t*) = *ηλ*_max_(*t*)*/*2 as a *warning signal*, not a safety certificate. Sustained *ρ*(*t*) *>* 1 indicates locally unstable updates along the sharpest measured direction and motivates stress testing for perturbation sensitivity under shift. Consequently, *ρ*(*t*) and *T*_EoS_ = Pr[*ρ*(*t*) *>* 1] should be reported as *hazard-linked training diagnostics* only when they are connected to a stated robustness or optimization-stability claim, with their assumptions validated against domain-relevant stress-test degradation (Section 6).

The hazard is not curvature itself, but making robustness claims without checking whether high-curvature training behavior corresponds to perturbation sensitivity. Curvature-aware objectives (e.g. Sharpness-Aware Minimization [SAM]) and schedule interventions can reduce sharpness in some settings, but they should be treated as candidate interventions whose value must be demonstrated under the same stress tests, not as causal proof of clinical safety.

### 3.1 Curvature-Aware Optimization as a Conditional Intervention

Conventional optimisers such as SGD, Momentum, Adam, and AdamW remain necessary baselines in stability-first biomedical ML. They differ in convergence speed, memory use, tuning sensitivity, and implicit regularisation, so optimiser choice can change both IID performance and behaviour under shift [32–34]. For this reason, optimiser comparisons should not report peak accuracy alone. They should state the learning-rate schedule, stopping rule, tuning budget, hardware, time-to-target, and degradation under domain-relevant stress tests.

Curvature-aware methods such as Sharpness-Aware Minimisation (SAM) and Gradient-Centralised SAM (GCSAM) [35, 36] should be treated as conditional interventions. They are most relevant when a paper makes optimisation-stability or robustness-under-shift claims, and when diagnostics such as *ρ*(*t*) or *T*_EoS_ suggest sustained high-curvature training. SAM-style objectives can reduce neighbourhood sensitivity in some regimes, but they also add computational cost, introduce extra hyperparameters, and may reduce performance under fixed budgets or heterogeneous biomedical data. Their value must therefore be demonstrated against strong conventional baselines under matched compute, matched tuning effort, and the same stress-test suite.

The CHB-MIT result illustrates this conditional logic. SAM with SGD underperformed SGD with Momentum on IID AUC (70.22% vs. 79.04%; Table 2), despite being motivated by stability. This negative result supports the reporting-first position: curvature-aware optimisation is not a default route to safer biomedical ML. It is a candidate intervention whose benefit must be verified through stress-test degradation, runtime cost, and sensitivity to tuning.

Across CIFAR-10 [37] and CHB-MIT [38], optimiser choice changes both predictive performance and time-to-target; see Table 2 and Fig. 5. These results motivate compute-aware reporting. A stability-first optimiser comparison should place each method on an efficiency-reliability frontier: IID

**Figure 4:**
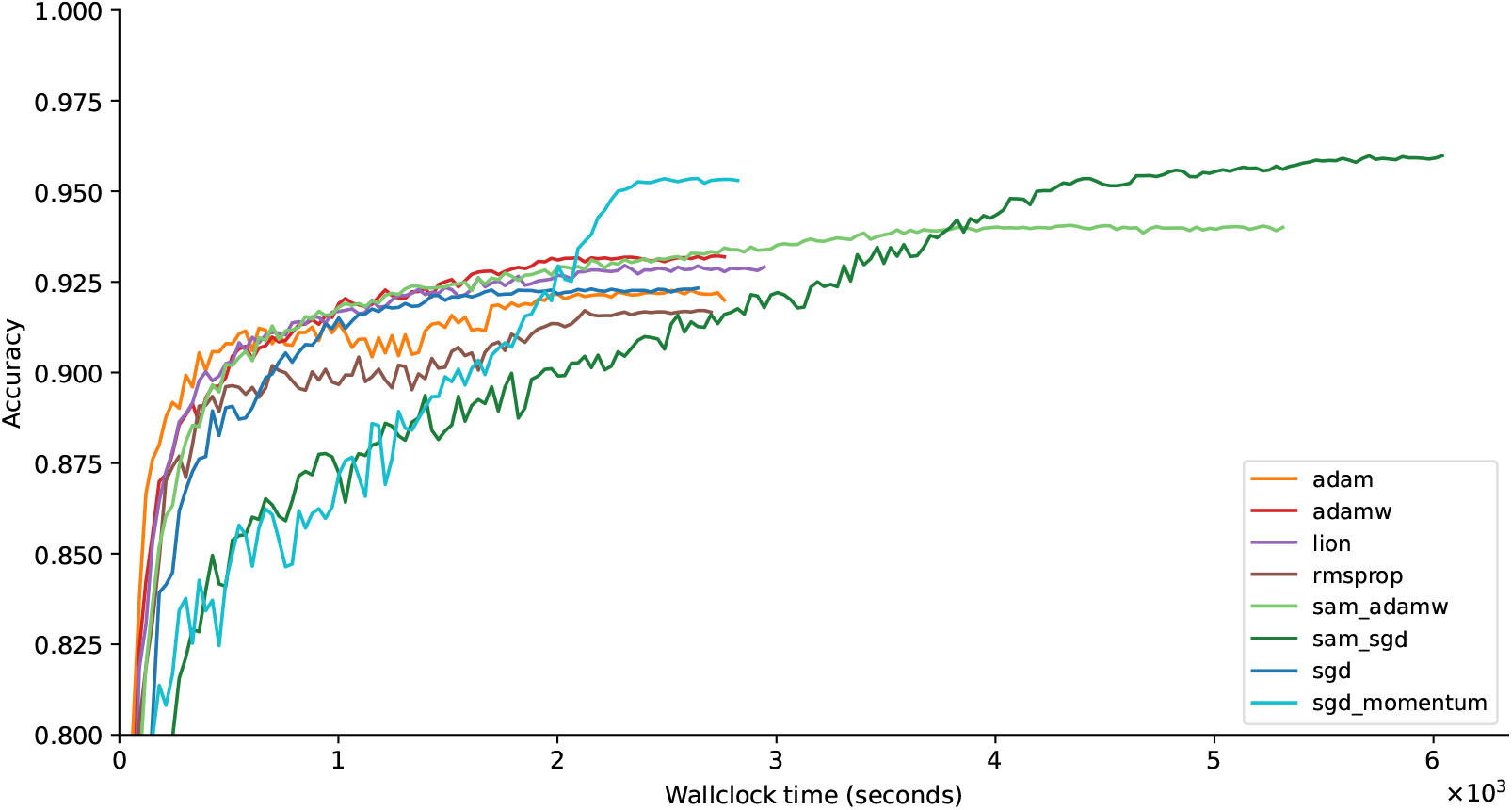
CIFAR-10 test accuracy as a function of wall-clock training time. Most methods achieve rapid gains within the first few hundred seconds, then show incremental improvements. Adam and AdamW yield fast initial gains, whereas SAM variants improve more slowly and can reach higher final accuracies with longer time budgets.

**Figure 5:**
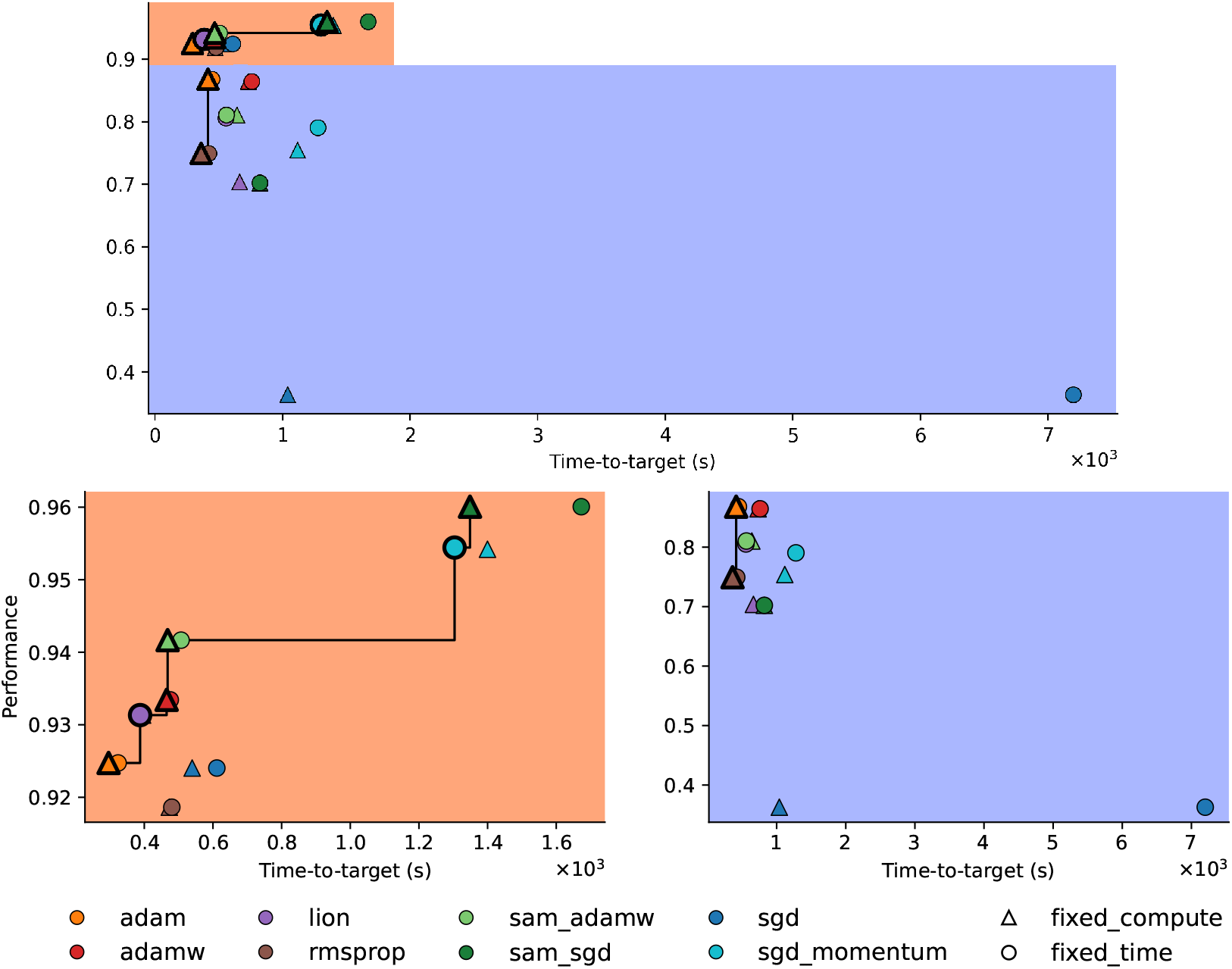
Efficiency and performance trade-off. The top panel overlays CIFAR-10 (*orange* background) and CHB-MIT (*blue* background) results in an efficiency and performance plane. The bottom row separates CIFAR-10 (IID test accuracy) and CHB-MIT (IID test AUC). The horizontal axis measures time-to-target in seconds (right-censored at the fixed-time budget when the target is not reached); accordingly, CHB-MIT SGD points lie on a budget-censored tail and should be interpreted as failures-to-target rather than unusually slow convergence. Pareto-optimal configurations are emphasized with thicker outlines and connected by a stepwise frontier.

## 4 Alternative Views and Objections

The following section outlines potential arguments against the claims in this paper and explains how we address these concerns in our call to action.

### Objection 1: Stability-first methods are too expensive

As mentioned in Section 3.1, NODEs and SAM-style objectives can increase computation. NODEs require numerical integration [39], and SAM adds extra forward/backward passes and can worsen the time-to-target under fixed budgets [35, 40]. In many biomedical workflows (edge or closed-loop), latency, memory, and power constraints are decisive, so strong discrete-time baselines may be the only viable choice [41]. **We therefore argue for *diagnosis-first* rather than “NODE+SAM by default”**: require irregular-time stress tests and hazard-linked training diagnostics, and escalate to continuous-time or curvature-aware interventions only when robustness curves and/or sustained *T*_EoS_ justify the cost (Sections 7 and 9).

### Objection 2: Discrete-time deep learning is sufficient if augmentation is strong

A common view is that corruption augmentation and data deficit modeling can substitute for continuous-time structure [22, 42]. We agree that strong discrete baselines can be sufficient when sampling is regular, horizons are short, and robustness curves are flat; our compute-frontier guidance shows this (Section 8). Our claim is narrower: when observation schedules vary materially and long-horizon stability is required, irregular-time stress tests often reveal failure modes that IID evaluation misses. In many biomedical settings, heavy augmentation is a response to limited datasets that are often constrained due to privacy. This can make “robustness” reflect researcher-chosen corruptions rather than real deployment shift. **We recommend releasing privacy-preserving, multi-site evaluation benchmarks, so claims about training stability and robustness can be independently verified beyond single-site IID splits**.

### Objection 3: Curvature/EoS metrics are not causally tied to deployment failures

We do not treat *ρ*(*t*) or *T*_EoS_ as safety certificates. **Our position is that these metrics are useful *hazard-linked diagnostics* only insofar as they predict stress-test degradation (Section 6)**. If correlation is weak for a task, the diagnostic should be deprioritized.

### Objection 4: Regime shift can be handled with fine-tuning or Test-Time Adaptation (TTA)

Shift-mitigation methods such as fine-tuning, domain adaptation, or test-time adaptation aim to recover performance after deployment shift is detected. **Our proposal differs by treats shift sensitivity and optimization instability as pre-deployment hazards that must be verified by stress tests and hazard-linked diagnostics**. Adaptation can be a valid mitigation, but it introduces additional risks [43, 44], and should not substitute reporting shift sensitivity curves.

## 5 Design Targets for Stability-First Biomedical AI

A stability-first approach is not a single model class or optimizer. So far, we narrow the required standard to three auditable targets: robustness under irregular observation, validity of any training-stability diagnostic used to support robustness claims, and task-appropriate early-warning evaluation when the task is transition-driven. Other considerations, including mechanistic plausibility and deployment constraints, remain important in some biomedical settings but are treated as optional extensions rather than core requirements.

### Target 1. *A stability-first report should quantify performance degradation under irregular observation schedules*

For forecasting, stability requires that error does not diverge when observation schedules change. The controlled test in Fig. 3 provides one concrete check, with protocol details in Appendix A.2. A biomedical analogue is event forecasting from EEG or Intensive Care Unit (ICU) records, where sampling patterns vary with workflow and acuity. The objective is to report behavior under schedule perturbations, not only high IID accuracy.

### Target 2. *A stability-first report should make training-stability diagnostics explicit, conditional, and stress-test linked*

EoS behavior shows that decreasing loss need not imply stable local geometry. Tracking *λ*_max_ during training can diagnose whether optimization operates in a locally high-curvature regime, but this is not a safety certificate or a default prescription for curvature-aware optimization. The measurement procedure used here is given in Appendix A.1, and the associated stability bound is derived in Appendix D.1. Diagnostics such as *ρ*(*t*) and *T*_EoS_ should be reported only with their assumptions and their empirical relationship to domain-relevant stress-test degradation.

### Target 3. *A stability-first report should use task-appropriate early-warning metrics for transition-driven biomedical problems*

For transition-driven tasks, such as seizure forecasting or acute deterioration prediction, evaluation should not rely only on classification accuracy. Reports should include lead time at a fixed false alarm rate where applicable, along with calibration of the warning signal and a clear definition of the pre-event window.

### Optional extensions

Mechanistic plausibility checks and deployment reporting remain valuable when the paper makes generative, edge, or closed-loop claims. In imaging, plausibility checks may include anatomical topology or intensity constraints; in physiological forecasting, they may include boundedness or non-negativity; and in edge settings, they may include latency, memory, energy, and activity sparsity. We therefore keep these topics as appendix-supported extensions rather than required pillars of the main reporting standard.

### 5.1 Audit criteria for the targets

In this section, we provide target guidance and specify what evidence would satisfy each target or falsify it.

**Target 1:** Report performance as a function of observation perturbations, including thinning rate, missingness, and time jitter. When forecasting is required, report whether long-horizon error remains bounded under schedule perturbation. A controlled construction is provided in Appendix A.2.

**Target 2:** Report a curvature proxy during training, such as *λ*_max_, alongside the learning-rate schedule. Report whether training enters an EoS regime (for example, whether *λ*_max_ exceeds 2*/η* for sustained periods) and whether curvature-aware optimization reduces time spent above this boundary. The report should also state the diagnostic-validity link: which deployment-relevant stress test the diagnostic is expected to predict, what direction of association is expected, and what result would weaken or falsify the diagnostic. See Appendices A.1, D.1 and D.2.

**Target 3:** For transition-driven tasks, report lead time at a fixed false alarm rate, define the pre-event window, and state how alarms outside that window are counted. If latent monitoring signals are used, report their calibration and whether they improve lead time over classification-only baselines.

### Optional extensions

For papers making generative, edge, or closed-loop claims, authors should state any additional plausibility or deployment checks matched to the modality. In imaging, this may include anatomical topology constraints or intensity-boundedness checks; in physiological forecasting, this may include boundedness or non-negativity; and in edge settings, this may include latency, memory, energy, and activity sparsity. These checks are optional extensions to the core reporting standard, not required targets for every biomedical ML paper.

The experiments in this paper instantiate the required reporting targets in Targets 1 to 3. Mechanistic plausibility and deployment checks are treated as optional extensions when the corresponding claims are made, with compact templates provided in the appendix.

## 6 Predictions and Falsification Tests

Position papers are only useful if they can be wrong. We therefore state falsifiable claims implied by the stability-first design approach. These predictions are stated as falsifiable hypotheses and accompanied by recommended minimal validation studies. Where only illustrative demonstrations are shown (e.g. FitzHugh-Nagumo [45]; PathMNIST [46]), we treat them as mechanism probes rather than domain-complete evidence: see Fig. 3 and Appendix A.2 for the FitzHugh-Nagumo experiment, and Fig. 2 and Appendix A.1 for the Edge-of-Stability experiment.

### Prediction 1. *Time-aware models are more robust to irregular resampling than imputation-based discrete baselines, but this advantage can vanish below the noise breakpoint* SNR^*⋆*^

We test this by schedule-perturbation stress tests (Fig. 3; protocol in Appendix A.2) and an SNR sweep that reports both performance and rollout-error boundedness; SNR^*⋆*^ is the smallest SNR where the continuous-time model’s degradation slope exceeds a discretely regularized baseline by a pre-specified margin (e.g. additional ΔAUC drop *>* 2% under matched budgets).

### Prediction 2. *At matched IID performance, lower T*_EoS_ *predicts smaller degradation under domain-relevant stress tests*

We operationalize this by reporting *T*_EoS_ alongside performance-versus-corruption curves and testing whether *T*_EoS_ correlates with degradation across seeds/configurations. This prediction is supported only if the direction of the association and the relevant stress test are specified before evaluation; it is weakened or falsified if *T*_EoS_ does not predict degradation under the stated perturbation at matched IID performance.

### Prediction 3. *For transition-driven tasks, latent monitoring improves lead time at a fixed false alarm rate compared with classification baselines*

We evaluate this by lead time at a fixed false alarm rate and, when applicable, by whether the warning signal co-varies with CSD proxies.

### Several evaluation metrics are provided for each prediction

For Prediction 1, report performance sensitivity to thinning and time jitter and report long-horizon error growth. For Prediction 2, report IID and stress-test performance alongside *T*_EoS_ (or area-under-*ρ*(*t*) above 1) and whether it correlates with stress-test degradation across seeds/configurations. For Prediction 3, report lead time at fixed false alarm rate and calibration of early warnings. For generative reconstruction or deployment claims, report optional plausibility and runtime checks using the appendix templates.

## 7 Minimum Reporting Standard

Safety-relevant biomedical ML claims should extend beyond IID metrics. Under tight space constraints, we propose two *required* pillars that directly audit the hazards in Positions 1 and 2. For transition-driven problems, the same reporting logic extends to early-warning evaluation: authors should report lead time at a fixed false alarm rate, define the pre-event window, and state how alarms outside that window are counted. Optional add-ons are reported in the appendix when relevant. Robustness should be structured and auditable, not ad hoc. This is supported by *Xian et al*. [47], who calls for standardizing robustness assessment via specification-driven testing by defining degradation mechanism, task priorities and then evaluating them accordingly.

### Required (P1) Irregular-time stress tests

Report performance as a function of observation perturbations, including thinning, timestamp jitter, bursty missingness, channel dropout, and SNR shift, and summarize degradation slopes, not only single-point scores. Where forecasting is claimed, report whether long-horizon error remains bounded under schedule perturbations.

### Required (P2) Hazard-linked training stability diagnostics

When training-stability or robustness claims are made, report at least one relevant training-stability metric (e.g. *ρ*(*t*) = *ηλ*_max_(*t*)*/*2 and *T*_EoS_), state the operational hazard it is intended to detect, and evaluate that hazard under at least one domain-relevant stress test with an explicit acceptance criterion (e.g. AUC drop *<* 5% under 30% channel dropout).

### Diagnostic-validity requirement

A training-stability diagnostic is valid for a given claim only if its operational interpretation is specified before evaluation. Authors should report: (i) the diagnostic being measured, (ii) the hypothesised hazard it is intended to detect, (iii) the stress test used to evaluate that hazard, (iv) the acceptance or failure threshold, and (v) the decision rule linking the diagnostic to the claim. For example, reporting *T*_EoS_ is insufficient unless the paper also states whether higher *T*_EoS_ is expected to predict greater degradation under a specified perturbation, such as channel dropout, SNR shift, or schedule jitter.

A stability-first report is non-compliant if it (i) omits schedule-perturbation sensitivity curves and (ii) uses training-stability diagnostics without tying them to an explicit stress-test acceptance bound (for example, AUC drop *<* 5% under 30% channel dropout or RMSE increase *<* 10% under 2 *×*thinning). For transition-driven tasks, lead-time and false-alarm-rate definitions should be reported using the same task-specific threshold logic. These thresholds are task-specific and should be justified by clinical context and failure cost.

Calibration/abstention reporting, mechanistic plausibility checks for generative models, and deployment reporting (latency/memory/energy on target hardware) remain optional add-ons for papers making high-stakes, generative, or closed-loop claims; we provide compact templates in the appendix.

## 8 Failure modes and limits of stability-first design

Although stability-first reporting can improve biomedical ML evaluation, it should not be read as a universal preference for more complex models. Continuous-time integration can amplify high-frequency measurement noise when the latent signal is weak, and curvature-aware optimization can be compute-intensive and sensitive to hyperparameters (e.g. SAM radius), with gains attenuating under tight budgets. Mechanistic plausibility constraints and deployment checks are important when those claims are made, but they are not required for every biomedical ML paper. Finally, when sampling is regular and dynamics are well-captured by discrete representations, strong discrete-time baselines may remain preferable on efficiency grounds.

The following compute-frontier heuristics summarize when the added cost of continuous-time modeling or curvature-aware optimization is most likely to be justified.

- **Prefer discrete-time baselines** when sampling is regular, horizons are short, and stress-test degradation is small (flat robustness curves), or when the signal is noise-dominated.
- **Prefer continuous-time / principled discretization (NODE/SSM)** when observation schedules vary materially across sites or workflows and long-horizon stability under schedule perturbations is a stated requirement.
- **Prefer curvature-aware interventions (e.g. SAM fine-tuning)** when training exhibits sustained *T*_EoS_ and robustness curves show large degradation slopes under clinically relevant corruptions.

### Stability–performance trade-off

Stability interventions (e.g., continuous-time solvers, SAM-style objectives, plausibility constraints) often trade compute and time-to-target for flatter solutions and reduced stress-test degradation, and may not improve peak IID performance under fixed budgets. We therefore treat stability as an explicit design objective with measurable cost: models should be compared on an efficiency–reliability frontier (IID score, degradation slope under shift, and compute), and escalated to higher stability tiers only when stress tests indicate clinically meaningful hazard.

## 9 Practical Adoption: Tiered Stability-First Compliance

We acknowledge that stability-oriented evaluation and intervention can introduce non-trivial computational costs. The reporting standard should therefore be adopted as an escalation pathway rather than an all-or-nothing mandate. We propose three tiers: **T1 (cheap): diagnosis under stress, T2 (moderate compute): stabilize optimization when diagnostics justify it**, and **T3 (high compute, high stakes): continuous-time or deployment-specific reporting when the use case requires it**. Higher levels subsume lower-tier requirements and justify correspondingly stronger claims.

### Tiered adoption and claim guardrails

Stability-first practices do not need to be adopted all at once. We recommend stating a compliance tier alongside the paper’s claims to prevent over-claiming: T1 supports evaluation-centric robustness characterization; T2 supports optimization-stability claims using *ρ*(*t*) / *T*_EoS_ related to stress tests; T3 supports deployment-oriented claims and requires on-target reporting. The expanded requirements and templates are deferred to Appendix C.

### Compute-aware implementation guidance

Ranking enables incremental adoption without requiring maximal compute from the outset. T1 introduces minimal overhead, primarily stress tests and calibration reporting. T2 concentrates additional compute where it yields the largest stability benefit, for example, by applying SAM-style objectives only during a final fine-tuning phase. T3 reserves solver-heavy continuous-time modeling and full deployment reporting for scenarios where clinical stakes justify the cost, for example, closed-loop or edge operation.

To reduce subjectivity, we recommend stating explicit acceptance thresholds that match the claimed use case. These thresholds are not universal; they should be justified by clinical context and failure cost.

## 10 Conclusion

Biomedical AI is increasingly deployed where failure is costly, but IID performance alone cannot establish reliability under irregular observation, missingness, and deployment shift. The central need is not a single model class, but auditable evidence that a system remains robust under deployment-relevant perturbations. We therefore argue for a minimum reporting standard: **(1) report irregular-time stress tests** with degradation slopes under thinning, jitter, data gaps, channel dropout, and SNR sweeps; **(2) make training-stability claims auditable** by reporting diagnostics such as *ρ*(*t*) and *T*_EoS_ only with their assumptions and validated links to stress-test degradation; **(3) use task-appropriate shift and early-warning metrics**, including lead time at a fixed false-alarm rate for transition-driven settings; and **(4) adopt tiered escalation rather than default complexity**. Strong discrete-time baselines may be sufficient when stress-test curves are flat. Continuous-time models, curvature-aware optimization, and deployment-specific reporting should be escalations justified by evidence, not default prescriptions. This reporting-first framing makes biomedical ML robustness claims more transparent, falsifiable, and clinically accountable.

## Data Availability

All datasets analysed in the present study are publicly available open-access resources. The CHB-MIT Scalp EEG Database is available via PhysioNet at https://physionet.org/content/chbmit/1.0.0/. The PathMNIST and CIFAR-10 datasets are also openly accessible via their respective original repositories. No new primary datasets were generated for this study.

https://medmnist.com/

https://www.cs.toronto.edu/~kriz/cifar.html

## Acknowledgments and Disclosure of Funding

The authors acknowledge the support from the Australian Research Council under Project DP230100010. We also thank the School of Biomedical Engineering at the University of Sydney for providing the computational resources required for this work.

## Author Contributions

Z.A.Z. conceptualised the study, developed the methodology, and drafted the manuscript. O.K. provided overarching supervision, funding acquisition, and strategic direction. I.A., L.F.H.C., and Z.H. contributed to the experimental design, assisted with the data analysis, and critically revised the manuscript for intellectual content. All authors reviewed and approved the final submission.

## Appendix A Experiments

### A.0.1 Compute resources and software

All experiments were run on a single Linux workstation (1*×* NVIDIA GB10 GPU; ARM aarch64 20-core CPU; 119 GiB RAM). We used fixed random seeds per run and report wall-clock runtime aggregated across seeds. Peak GPU memory was recorded via `torch.cuda.max memory allocated()` and `torch.cuda.max memory reserved()`.

Software: Python 3.12.3, PyTorch 2.11.0.dev20260212+cu130, CUDA 13.0, cuDNN 91701.

### A.0.2 Data and licenses

All datasets used in this study are publicly available open-access resources and were used in accordance with their respective licenses: the CHB-MIT Scalp EEG Database under the Open Data Commons Attribution License v1.0, PathMNIST under the Creative Commons Attribution 4.0 International (CC BY 4.0) license, and CIFAR-10 under the MIT license. We comply fully with all attribution and redistribution terms.

### A.1 Optimization Instability (Edge of Stability)

We quantify optimization instability in the EoS regime by jointly tracking (i) training loss and (ii) a curvature-based sharpness proxy: the highest eigenvalue of the Hessian parameter of a fixed batch loss during SGD training of a Multilayer Perceptron (MLP) on PathMNIST [46]. Since the estimate is computed on a fixed mini-batch rather than the full training objective, the aforementioned proxy captures local curvature rather than the full training objective. The analysis uses the classical quadratic base stability condition for gradient descent as a reference threshold and evaluates whether the measured curvature persistently exceeds it, as is commonly done in EoS studies [11, 30]. We report loss and sharpness trajectories, along with a 2D loss landscape slice and a projected optimization path (Fig. 2). A summary of the training protocol and aggregate statistics is given in Table 1. We are training a non-linear neural network with mini-batch SGD, so the Hessian is not guaranteed to be positive nor stable, and batch noise can cause brief spikes in the estimated curvature. Using SGD, we interpret 2*/η* only as a diagnostic baseline, not a convergence guarantee. The purpose is to report and compare the observed curvature relative to a widely used theoretical reference [11, 30].

**Table 1:** Summary statistics for the Edge-of-Stability experiment (10 seeds), visualized with Fig. 2.

|  |  |
| --- | --- |
| **Training setup** |  |
| Architecture | 3-layer MLP, 2352→256→128→9 |
| Optimizer | SGD (no momentum) |
| Learning rate $\eta$ | 0.01 |
| Batch size $B$ | 128 |
| Epochs $T$ | 50 |
| **Landscape** |  |
| PCA variance | PC1: 89.1%, PC2: 6.8% |
| **Metrics (n=10)** |  |
| Stability limit | $2/\eta = 200.00$ |
| Peak sharpness | $382.5 \pm 63.0$ |
| Peak sharpness range | $[306.9, 522.0]$ |
| Final sharpness | $227.5 \pm 38.1$ |
| Initial loss | $1.824 \pm 0.011$ |
| Final loss | $0.784 \pm 0.009$ |
| Loss reduction | 57.0% |

**Table 2:** End-to-end performance and efficiency benchmarks (fixed-time, 2 h). Bold indicates the highest predictive performance and the fastest convergence time for each dataset. We report mean *±*s.d. over *n* = 10 seeds. Statistical comparisons use two-sided Wilcoxon rank-sum tests for matched pairs (SGD *vs*. SAM (SGD); AdamW *vs*. SAM (AdamW)), with Holm-Bonferroni correction. Time (s) denotes time-to-target (Acc/AUC at 0.90) when reached; otherwise it is right-censored at the 2 h budget (7200 s).

| Optimizer | CIFAR-10 |  | CHB-MIT |  |
| --- | --- | --- | --- | --- |
| | Acc. (%) (↑) | Time (s) (↓) | AUC (↑) (%) | Time (s) (↓) |
| SGD | 92.40 ± 0.19 | 539 ± 105 | **36.27 ± 14.99** | **7200 ± 0** |
| SGD + Momentum | 95.44 ± 0.08 | 1304 ± 136 | 79.04 ± 2.55 | 1279 ± 507 |
| Adam | 92.47 ± 0.12 | **296 ± 19** | **86.80 ± 0.55** | 415 ± 181 |
| AdamW | 93.34 ± 0.07 | 465 ± 45 | 86.45 ± 0.29 | 732 ± 546 |
| RMSProp | 91.86 ± 0.06 | 473 ± 138 | 74.93 ± 21.59 | **362 ± 60** |
| Lion | 93.13 ± 0.09 | 395 ± 48 | 80.54 ± 3.48 | 556 ± 140 |
| SAM (SGD) | **96.01 ± 0.03<sup>†</sup>** | 1349 ± 233 | 70.22 ± 12.88 | 822 ± 7 |
| SAM (AdamW) | 94.17 ± 0.12<sup>†</sup> | 468 ± 10 | 81.03 ± 9.98<sup>†</sup> | 641 ± 116 |

<sup>†</sup> Indicates a statistically significant improvement over the matched baseline after Holm-Bonferroni correction.
Red indicates right-censoring: CHB-MIT SGD did not complete an epoch within 2 h; AUC is the last available evaluation and time is set to 7200 s.

We train a fully connected MLP (2352*→* 256 *→* 128*→* 9) on PathMNIST [46] using SGD [48] with loss of cross-entropy, batch size *B* = 128 and *T* = 50 epochs. PathMNIST is a dataset of 28 *×*28-pixel patches of slides stained with hematoxylin and eosin (H&E staining), and we input the dataset into 3 channels; therefore, the flattened input is fed into 28*×* 28*×* 3 = 2352 neurons. The data set consists of 9 unique tissue types, and the MLP is trained to classify them (output layer with 9 neurons) [46]. This configuration is chosen as a controlled, computationally lightweight setting in which curvature can be estimated at every epoch, and results can be replicated across many random seeds. We fix the learning rate at *η* = 0.01 to intentionally probe the EoS regime, using the classical quadratic-basin stability reference *λ*_max_ *≤* 2*/η* from gradient descent theory [49, 50] as a diagnostic baseline (rather than a convergence guaranty). All results are aggregated across the seeds *S* = 10, with confidence intervals, to demonstrate robustness to random initialization and batch order. Each seed fixes all pseudo-random generators (NumPy, PyTorch CPU, and PyTorch CUDA when available), reinitializes model parameters, and reshuffles batches. Per epoch, we record the mean training loss in all batches and an estimate 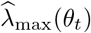 of the largest Hessian eigenvalue of the (probe-batch) loss in the current parameters *θ*_*t*_. We aggregated the results between the seeds with the mean between the seeds and a normal approximation of 95% confidence interval 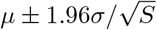.

#### Reference stability condition and the 2*/η* threshold

Let *ℒ* : ℝ^*d*^*→* ℝ denote the loss objective and consider a full-batch gradient descent

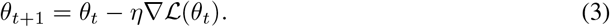

To connect curvature and stability, the standard local quadratic approximation used in optimization texts is used [49, 50]. Around a stationary point *θ*^*⋆*^ with ∇ℒ (*θ*^*⋆*^) = 0, the linearization of the gradient yields

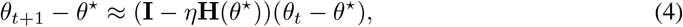

where **H**(*θ*^*⋆*^) = ∇^2^ℒ (*θ*^*⋆*^) is the Hessian. A sufficient and essentially tight condition for local convergence of Eq. (4) is

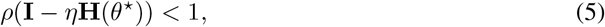

where *ρ*(*·*) denotes the spectral radius. In a convex quadratic basin, **H**(*θ*^*⋆*^) is symmetric positive semidefinite with eigenvalues 0*≤ λ*_1_*≤··· ≤ λ*_max_, so the eigenvalues of **I***− η***H** are (1 *− ηλ*_*i*_) and the condition reduces to

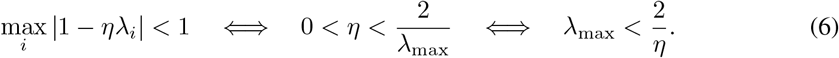

We therefore use the nominal threshold 2*/η* = 200.0 (for *η* = 0.01) as a reference line in our sharpness plots [49, 50]. Following prior EoS analyzes [11, 30], we interpret sustained violations of Eq. (6) by 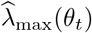 as evidence that training operates outside the locally stable quadratic regime. Eq. (6) is exact for quadratic objectives under full-batch gradient descent. Our setting involves non-convex networks and SGD, so **H**(*θ*_*t*_) may be indefinite and stochasticity can induce transient excursions. Accordingly, we treat 2*/η* as a diagnostic baseline, not as a guarantee of divergence or convergence. The purpose is to report and compare observed curvature relative to a widely used theoretical reference [11, 30].

We estimate *λ*_max_(**H**(*θ*_*t*_)) by iterating the power on a fixed probe batch (*x, y*). Let *l*(*θ*; *x, y*) denote the probe-batch loss, with gradient *g*(*θ*) = *∇*_*θ*_*l*(*θ*; *x, y*) and Hessian 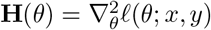. Starting from a random unit vector *v*_0_, the power iteration updates

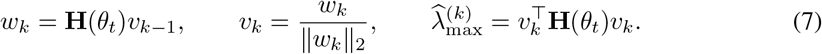

We compute *w*_*k*_ = **H***v*_*k−*1_ without forming **H** using Hessian-vector products (HVPs),

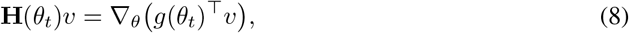

implemented via automatic differentiation. We run at most *K* = 50 iterations with tolerance 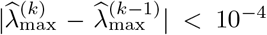. The probe batch is constructed deterministically (fixed indices, no shuffling) and is held constant across epochs within a run; we store the probe batch with each checkpoint to ensure exact reproducibility of the reported sharpness trajectories.

For each seed, we save (i) per-epoch metrics, (ii) model parameters (state dict), (iii) the fixed probe batch used for curvature estimation, and (iv) epoch-level parameter snapshots used for the loss-landscape visualizations. All figures are generated solely from these saved artifacts.

#### A.2 Topological Stability and Irregular Sampling (FitzHugh-Nagumo)

We benchmarked on FitzHugh-Nagumo neuronal dynamics [45, 51] with parameters (*I*_ext_ = 0.5, *a* = 0.7, *b* = 0.8, *τ* = 12.5). We compared a Neural ODE [52] against two baselines designed for irregularly sampled sequences. Crucially, all models were trained on the exact same supervised, state-to-state forecasting task: given the current observation **x**(*t*_*i*_) and the elapsed time Δ*t*_*i*_ = *t*_*i*+1_ *−t*_*i*_, predict the next state **x**(*t*_*i*+1_). This ensures a fair baseline comparison by matching the learning objective across architectures, with differences arising only from the model class and how irregular time is incorporated. **visualization determinism**. The 2D loss-slice shown in Fig. 2 is defined by an anchor checkpoint (*θ*_*c*_) and two directions (*d*_1_, *d*_2_) in parameter space, where (*d*_1_) is the dominant curvature direction (top Hessian eigenvector at *θ*_*c*_) and *d*_2_ is an orthogonal complement. Since the orthogonal complement is not identifiable (there are infinitely many valid choices), selecting *d*_2_ is necessarily a convention; we avoid subjective “plane selection” by using a neutral orthogonal choice and then storing the realized slice directions (*d*_1_, *d*_2_) with each run. Consequently, all visualizations are regenerated deterministically from saved artifacts (checkpoints, probe batch, and stored (*d*_1_, *d*_2_)), and differences across regenerations reflect changes in the underlying saved artifacts rather than re-sampling a new slice plane. When run on GPU, exact pixel-level determinism may additionally require deterministic backend settings; we therefore report the software stack (Section A.0.1) and provide fixed seeds and stored slice directions.

1. **Time-Aware LSTM (T-LSTM)** [53]. Following *Baytas et al*., T-LSTM explicitly uses the time elapsed between successive observations, Δ*t*_*i*_ = *t*_*i*_*− t*_*i−*1_, through a monotone non-increasing decay function *g*(Δ*t*_*i*_) that discounts the *short-term* component of the previous cell state via a learned subspace decomposition of the cell memory (as defined in [53]).We used a 2-layer T-LSTM with hidden size *H* = 128. In our implementation, the model processes (**x**(*t*_*i*_), Δ*t*_*i*_) at each step and is trained to minimize the one-step prediction error between its output and the ground-truth next state **x**(*t*_*i*+1_).
2. **ODE-RNN** [20]. Following Rubanova et al., ODE-RNN defines a continuous-time hidden state that evolves between observation times according to an ODE solver step, and is updated at observation times with a discrete recurrent update (we use a GRU update, consistent with [20]). The readout is trained with the same one-step supervised forecasting objective, i.e., predicting **x**(*t*_*i*+1_) from **x**(*t*_*i*_) and Δ*t*_*i*_.

We use the T-LSTM cell [53], including the elapsed time discount through the non-increasing monotone decay function *g*(Δ*t*). In our experiments, we instantiate this model as a *two-layer* T-LSTM encoder (with hidden size *H* = 128) to better capture the irregularly sampled FitzHugh-Nagumo trajectories, while still keeping the comparison fair and the total number of parameters within a commonly accepted range. This choice of depth is an architectural design decision for our benchmark and is not a claim attributed to [53]. To control for model capacity, we recommend reporting parameter counts for all models and including a parameter-matched discrete-time baseline (e.g., a wider/deeper T-LSTM, or an RNN baseline with Δ*t* concatenated to the input as in Eq. (1) of [20]).

##### A.2.1 Mathematical framework for irregular continuous dynamics

We use the FitzHugh-Nagumo/Bonhoeffer-van der Pol (BVP) class of two-dimensional excitable dynamics [45, 51]. In our experiment, we employ the common rescaled form

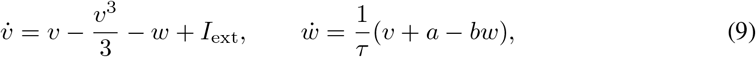

which, up to a change of variables, is equivalent to the BVP equations in [45]. We represent this as state **x**(*t*) = [*v*(*t*), *w*(*t*)]^*⊤*^ and parameters (*I*_ext_, *a, b, τ*) = (0.5, 0.7, 0.8, 12.5). We use this parameter configuration because it is a well-studied regime in which the system exhibits stable bounded oscillations (i.e., a limit-cycle attractor) [45, 51, 54].

For each training sequence, we sample an irregular observation schedule 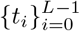with *t*_0_ = 0 and increments Δ*t*_*i*_ = *t*_*i*+1_ *− t*_*i*_ drawn from a uniformly jittered step size, Δ*t*_*i*_ = max(0.1, *µ*_Δ*t*_ + *ϵ*_*i*_) where *ϵ*_*i*_ ~ Unif(*−δ*_Δ*t*_, *δ*_Δ*t*_). We denote the resulting dataset by 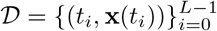.

###### Neural ODE training objective (matched to baselines)

Rather than training the Neural ODE by derivative matching, we train its vector field *f*_*θ*_ to solve the same supervised, one-step state forecasting task as the discrete-time baselines. For each observed pair (**x**(*t*_*i*_), Δ*t*_*i*_), we predict the next state by integrating the learned dynamics over the interval [*t*_*i*_, *t*_*i*+1_]:

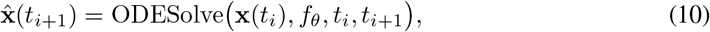

and we minimize the mean-squared one-step prediction error

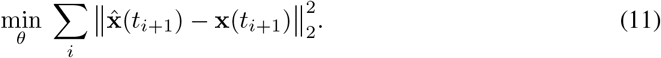

We backpropagate through the ODE solver (we use an explicit Runge-Kutta method, RK4) to optimize *θ*. At test time, we obtain rollouts on the grid {*t*_*i*_}y iterating the same integrate-and-predict step over each interval [*t*_*i*_, *t*_*i*+1_].

For completeness, our T-LSTM baseline appends the elapsed time as an additional input channel; that is, it processes (**x**(*t*_*i*_), Δ*t*_*i*_) at each step to predict the next state. Our ODE-RNN baseline follows the continuous-time hidden state construction of [20], evolving the hidden state between observations according to an ODE and applying a GRU update at observation times; in our implementation, the evolution between observations uses a single explicit Euler step *h← h* + Δ*tg*_*θ*_(*h*) followed by a GRUCell update with **x**(*t*_*i*_), and the readout is trained with the same one-step state forecasting loss.

#### A.3 End-to-End Optimizer Benchmark and Pareto Trade-off Plots (CIFAR-10 and CHB-MIT)

We benchmarked a set of end-to-end optimizers in two domains: CIFAR-10 image classification [37] and CHB-MIT EEG seizure detection [38, 55]. We benchmark a representative subset of the optimizers in Table 5 and report both IID performance and efficiency (time-to-target). For each dataset, we trained under fixed training-budget regimes and logged both (i) IID predictive performance and (ii) an efficiency cost. The resulting efficiency-performance points were aggregated in random seeds and visualized as Pareto trade-off plots (Fig. 5).

For CIFAR-10 [37], the main IID metric is the test accuracy. For CHB-MIT [38], the main IID metric is the test AUC, while the definition of the time-to-target uses the validation AUC as the gating metric (to avoid optimistic feedback from the test-set during training).

We consider two training-budget regimes: (i) *fixed time*, where training is stopped at a specified wall-clock budget; and (ii) *fixed compute*, where training is stopped at a specified compute proxy (for example, a fixed number of epochs or update steps). These regimes are shown as different marker shapes in the Pareto plots.

The efficiency cost is defined as the *time-to-target*, in seconds, namely the wall-clock time required to first reach a fixed target performance level (e.g., ACC at 0.90 for CIFAR-10; see Fig. 4, or AUC at 0.90 for CHB-MIT), when available. If time-to-target is not available for a run, we fall back to an aggregated wall-clock proxy derived from either the intended time budget or the empirically measured runtime. Configurations are excluded from the plot only when neither time-to-target nor a wall-clock proxy is available. The performance axis reports the aggregated IID metric (higher is better): test accuracy for CIFAR-10 and test AUC for CHB-MIT.

**Figure 6:**
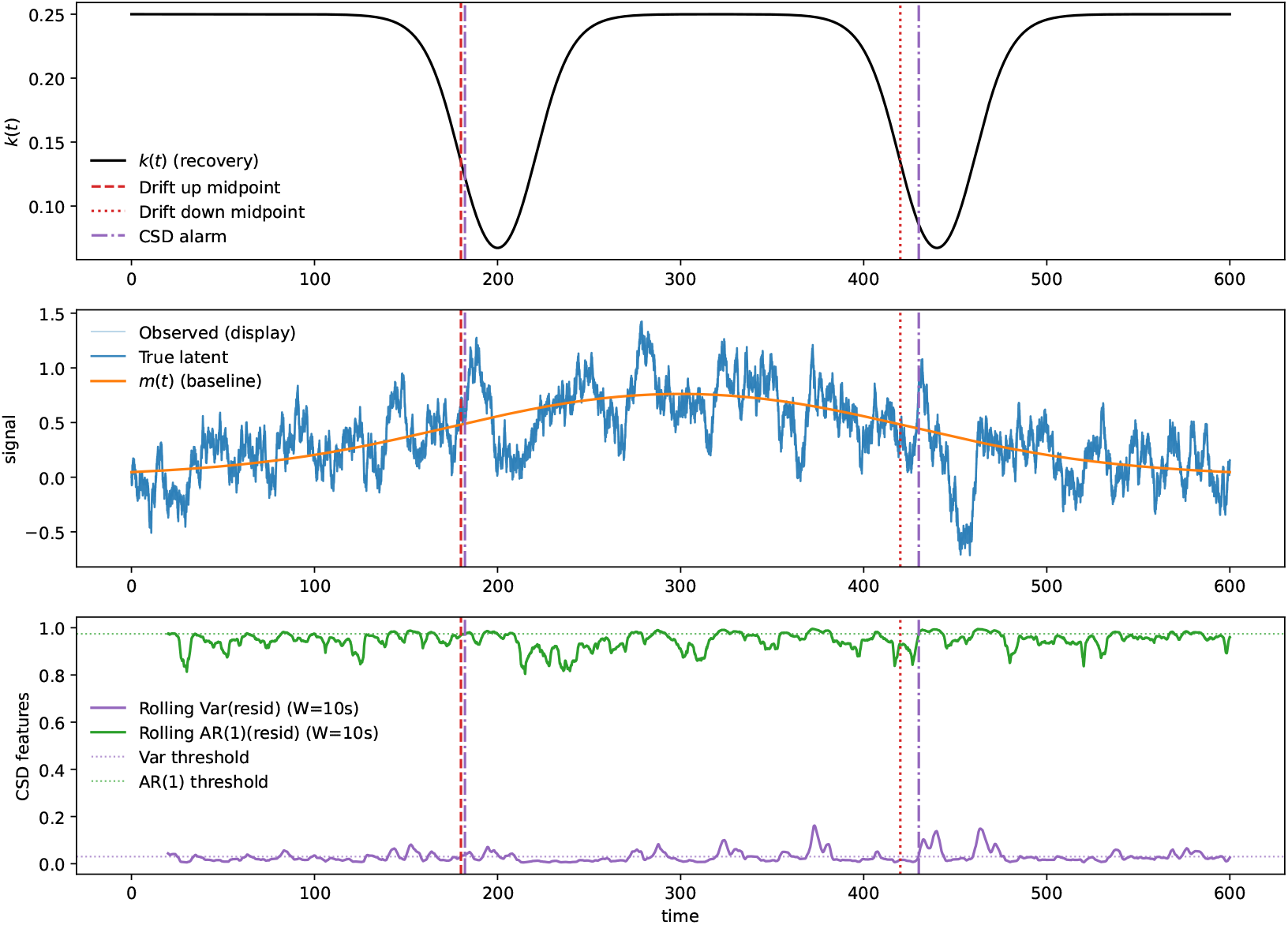
Two drift episodes with feature-only CSD alarms in a mean-reverting latent. Top: the recovery rate *k*(*t*) is held near a stable baseline but exhibits two local slow-down episodes (dips), aligned to an upward- and downward-drift episode (red vertical markers). Middle: the latent follows a slowly drifting baseline *m*(*t*) (orange), shown alongside the noisy observed trace used for monitoring. Bottom: rolling variance and lag-1 autocorrelation AR(1) are computed on detrended residuals and compared to thresholds calibrated on an early baseline window (horizontal dotted lines). A CSD alarm triggers when both AR(1) and variance exceed threshold for a sustained duration, with a cooldown to permit at most one alarm per episode (purple dash-dot markers).

Each configuration was evaluated in multiple random seeds (summary in Table 2). We aggregate each metric using the mean across successful runs, where a run is marked as successful if it yields a finite IID score. We additionally compute a failure rate as the fraction of runs that failed and encode it via marker transparency (a higher failure rate corresponds to more transparent markers).

On the CHB-MIT dataset, SGD is effectively right-censored by the 2 hour wall-clock limit, so the reported AUC reflects incomplete training rather than converged performance. Under tight compute budgets, plain SGD is not viable on CHB without momentum/adaptive scaling, so efficiency metrics are the meaningful comparison.

We treat the plot as a bi-objective optimization problem that minimizes efficiency cost (x) and maximizes performance (y). A configuration is Pareto-optimal if no other configuration is both faster (lower x) and better-performing (higher y). We compute the Pareto set and visualize it with thicker marker outlines and a stepwise frontier; see Fig. 5.

#### A.4 Critical Slowing Down Alarm Under Regime Shift

We provide a controlled mechanism probe demonstrating that standard early-warning statistics (rolling variance and lag-1 autocorrelation) can increase as a system’s recovery rate weakens, yielding an operational early-warning signal under distribution drift [56, 57]. The purpose is a sanity check for the monitoring protocol, not a claim that CSD universally holds in biomedical modalities.

##### Dynamical regime shifts

We simulate a 1D noise-driven relaxation process with a baseline *m*(*t*) that undergoes two regime shifts (upward drift followed by a return drift) and a time-varying recovery rate *k*(*t*):

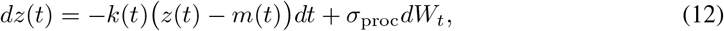

where *W*_*t*_ is a standard Wiener process. When *k*(*t*) is locally reduced, perturbations decay more slowly, which is the mechanism that produces CSD signatures [56]. Let the upward shift midpoint be *t*_↑_ and the downward (return) shift midpoint be *t*_↓_, with a shared smoothness parameter *w*_*m*_. We define

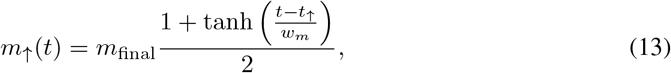

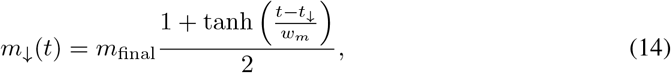

and take the net baseline as

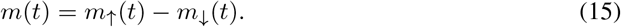

Thus, *m*(*t*) rises toward *m*_final_ and later returns toward 0, yielding two regime shifts in the mean level.

To align CSD behavior with each regime shift, we use two finite-duration slow-down episodes in *k*(*t*), each centered near (*t*_*↑*_, *t*_*↓*_). Concretely, *k*(*t*) stays near a baseline *k*_base_ but dips toward *k*_min_ in a local window around each shift, with dip sharpness controlled by a width parameter *w*_*k*_. This produces two periods of weakened recovery rather than a single global monotone ramp, supporting detection of two distinct alarm events.

##### Stochastic forcing and observation model

We simulate Eq. (12) using an Euler-Maruyama update with step size Δ*t* [58]. The observed latent is

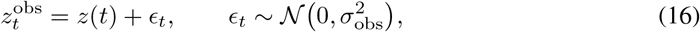

with *time-invariant* measurement noise. For visualization only, we optionally add slow baseline wander and apply a light Exponential Moving-Average (EMA) low-pass filter; critically, CSD statistics are computed on the unfiltered observation after rolling-mean detrending to avoid artificially inflating AR(1) via filtering or time-varying colored noise [57].

##### CSD monitoring statistics

We compute rolling-window early-warning statistics on detrended residuals [57]. For window length *W*, define the rolling mean 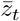 and residual

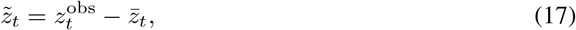

then report:

- Rolling variance: Var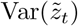 over the window.
- Rolling AR(1): the lag-1 autoregressive coefficient estimated within the window.

We interpret increasing AR(1) and variance as consistent with slowing recovery when *k*(*t*) is locally reduced (critical slowing down) [56, 57].

##### CSD alarm arming and cooldown

To turn early warning into a simple decision rule, we use a CSD alarm based on rolling AR(1) and variance. We set the alarm thresholds from an initial “baseline” segment of the trajectory using baseline z-score thresholds. Since AR(1) is naturally limited to the range [*−* 1, 1] [56], we apply a Fisher-*z* transform (that is, atanh(*·*)) when setting the AR(1) threshold so the calibration behaves sensibly and does not produce invalid cutoffs [59]:

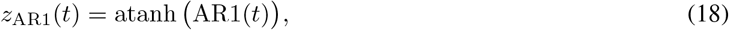

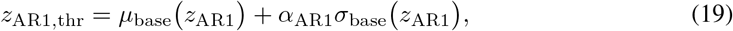

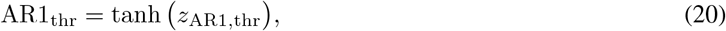

and we threshold variance in its native scale:

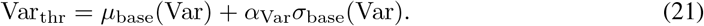

An alarm triggers at the earliest time when *both* AR1(*t*) *>* AR1_thr_ and Var(*t*) *>* Var_thr_ hold continuously for a sustained duration *τ* seconds. We additionally (i) *arm* the detector only when recovery is sufficiently weakened (i.e. *k*(*t*) *< k*_alarm_) to focus the alarm on the intended slow-down mechanism, and (ii) enforce a cooldown period after each trigger, allowing up to two alarms aligned to the two regime shifts. In the implementation, we optionally use different arming thresholds for the first and second alarm (*k*_alarm,1_, *k*_alarm,2_) to control relative timing.

##### A.4.1 Experimental setup

###### Parameters and reproducibility

We simulate a single trajectory of length *T* = 500 with step size Δ*t* = 0.01 (so *N* = *T/*Δ*t* samples) using a fixed random seed, which is long enough to contain a clean baseline segment plus two well-separated drift episodes while keeping the simulation lightweight. The baseline drift uses two regime-shift midpoints (*t*_↑_, *t*_↓_) = (220, 360) with width *w*_*m*_ = 120 and magnitude *m*_final_ = 1.0, chosen to create a clear mean shift that is gradual (drift-like) rather than an abrupt step. The recovery rate has baseline *k*_base_ = 0.25, minimum *k*_min_ = 0.01, and local slow-down width *w*_*k*_ = 20, yielding two finite-duration weakened-recovery episodes aligned to the two shifts (so two distinct CSD alarms are meaningful). Stochastic forcing is set to *σ*_proc_ = 0.15 to produce measurable fluctuations for estimating variance/AR(1), while observation noise is set smaller (*σ*_obs_ = 0.03) and time-invariant so changes in CSD features are attributable to recovery slow-down rather than a changing noise model. For visualization only, we optionally add slow baseline wander (amplitude 0.05 at 0.02 Hz) and apply an EMA low-pass filter with *α* = 0.05 to mimic biomedical acquisition effects; CSD statistics are computed on the unfiltered 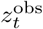after rolling-mean detrending to avoid artificially inflating AR(1).

###### Windowing and alarm configuration

We compute rolling AR(1) and variance using a *W* = 10s window, which smooths out moment-to-moment noise while still letting us see changes around each slow-down episode. We set the alarm thresholds from an initial baseline period (the first *f*_base_ = 0.10 of the run) so the cutoffs reflect “normal” pre-shift behavior. Thresholds are set using baseline z-score cutoffs, with *α*_AR1_ = 1.5 (applied in Fisher-*z* space) and *α*_Var_ = 1.0 (in variance space), so AR(1) is treated a bit more conservatively while variance remains sensitive enough to move during each episode. We trigger an alarm only if both features stay above threshold for *τ* = 2s, to avoid reacting to brief spikes. After an alarm fires, we wait 30s (cooldown) before allowing another alarm, so each episode produces at most one trigger. Finally, we only allow alarms when recovery is actually weakened (*k*(*t*) *< k*_alarm_); we use *k*_alarm,1_ = 0.18 for the first episode and a stricter *k*_alarm,2_ = 0.12 for the second episode so the second alarm does not fire too early.

###### Calibration statement

Unless otherwise stated, parameters not fixed by the simulation design were chosen via a small preliminary calibration on the baseline segment (first *f*_base_ fraction), with the objective of producing a stable false-alarm rate and visually interpretable alarms.

## Appendix B Evaluation Protocols and Reporting Templates

### B.1 Illustrative toy protocol: curvature vs generative stability

We outline a minimal falsification-style check for the hypothesis that sustained EoS exposure correlates with instability and mechanistic violations in score-based reconstruction. Train two matched models: **(A)** curvature-blind (standard optimizer/schedule) and **(B)** curvature-aware (e.g., SAM/GCSAM applied in a final fine-tuning phase). Log a training stability diagnostic (e.g., *ρ*(*t*) = *η*(*t*)*λ*_max_(*t*)*/*2 and *T*_EoS_ = Pr[*ρ*(*t*) *>* 1]) and compare the models under the same stress-test suite, reporting fidelity and plausibility-violation rates. This optional appendix protocol supports generative-reconstruction claims if model B matches fidelity while reducing instability or plausibility violations, especially under shift.

#### B.2 Mechanistic plausibility checks for generative reconstruction (metricised)

Pixel-level fidelity can miss clinically hazardous artifacts. We recommend reporting at least one mechanistic plausibility check as a falsification probe, alongside fidelity. Let 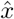denote the model output, *x*_ref_ a reference (when available), and *x*_base_ a conservative baseline reconstruction. See Table 3 for the compact set of plausibility checks.

**Table 3:**
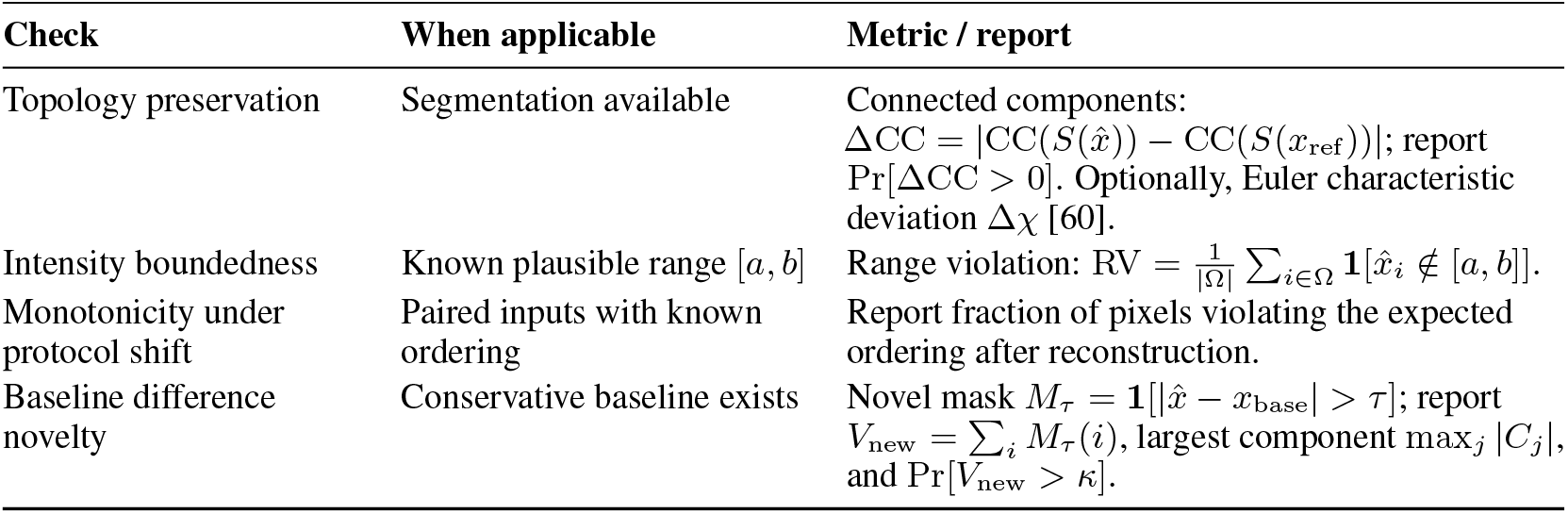
Compact plausibility checks for generative reconstruction. Report violation rates (not only means) and evaluate under stress.

**Table 4:** The geometry of health and disease: dynamical concepts for biomedical safety.

| Dynamical concept | Mathematical definition | Biomedical interpretation |
| --- | --- | --- |
| **Fixed point** | $\lim_{t \rightarrow \infty} x(t) = x^*$ | **Homeostasis.** A stable target state, such as regulated blood glucose [68, 69]. |
| **Limit cycle** | $x(t + T) = x(t)$ | **Biological rhythms.** Stable oscillations, such as the cardiac cycle [69, 70]. |
| **Chaotic attractor** | $\Lambda_{\max} > 0$, bounded | **Healthy variability.** Complex, non-repeating variability (for example, HRV) [71, 72]. |
| **Bifurcation** | Topological structural change | **Acute onset.** A sudden transition between clinical states (for example, seizure) [69, 73, 74]. |

**Table 5:** Optimiser families as conditional design choices for stability-first reporting. The table is not a ranking. Each optimiser can be appropriate depending on dataset size, compute limits, tuning budget, and the robustness claim being made. The reporting column states what must be disclosed when the optimiser is used to support stability or robustness claims. Abbreviations: EoS = Edge of Stability, LLM = Large Language Model.

| Optimiser | Family | Main role | Reporting relevance |
| --- | --- | --- | --- |
| SGD<br>[48] | Conventional | Simple stochastic gradient descent with low memory overhead. | Required baseline when feasible. Report learning-rate schedule, batch size, stopping rule, time-to-target, and stress-test degradation. |
| Momentum<br>[75] | Conventional | Adds a velocity term that can accelerate and smooth optimisation. | Strong conventional comparator for biomedical workloads. Report momentum coefficient, tuning budget, and whether gains persist under shift. |
| Adam<br>[76] | Adaptive | Uses first- and second-moment estimates to adapt per-parameter step sizes. | Useful when fast optimisation is needed. Report sensitivity to weight decay, learning rate, and whether adaptive scaling changes stress-test degradation. |
| AdamW<br>[33] | Adaptive | Decouples weight decay from adaptive step-size updates. | Strong adaptive baseline. Report weight decay, schedule, tuning budget, time-to-target, and stress-test deltas. |
| RMSProp<br>[77] | Adaptive | Uses a moving average of squared gradients for per-parameter scaling. | Relevant for non-stationary or noisy objectives. Report variance across seeds and whether apparent gains survive under perturbation tests. |
| Lion<br>[78] | Adaptive | Uses sign-based momentum updates with low memory use. | Relevant when memory or large-batch efficiency matters. Report stability across seeds, tuning sensitivity, and compute-normalised performance. |
| Adan<br>[79] | Adaptive | Incorporates Nesterov-style gradient difference information. | Useful as a fast-convergence comparator. Report whether faster convergence changes robustness or only IID performance. |
| LARS<br>[80] | Adaptive | Applies layer-wise adaptive rate scaling for large-batch training. | Relevant for large-batch regimes. Report batch size, scaling rule, hardware, and whether large-batch training increases sharpness or stress sensitivity. |
| Sophia<br>[81] | Second-order proxy | Uses low-cost curvature information to adapt updates. | Relevant when curvature information is part of the optimisation claim. Report the curvature proxy, update frequency, compute overhead, and stress-test deltas. |
| SAM<br>[35] | Curvature-aware | Minimises loss under neighbourhood perturbations to favour flatter solutions. | Candidate intervention when sharpness diagnostics suggest perturbation sensitivity. Report radius, extra compute, matched baseline, tuning effort, and stress-test degradation. |
| GCSAM<br>[36] | Curvature-aware | Combines SAM-style neighbourhood optimisation with gradient centralisation. | Candidate intervention for robustness claims. Report whether gains persist under matched compute and domain-relevant perturbations. |
| Muon<br>[82] | Matrix-based | Uses matrix-structured updates or orthogonalisation for scalable training. | Relevant for large models. Report memory overhead, hardware assumptions, tuning sensitivity, and whether benefits transfer to biomedical stress tests. |
| Kron<br>[83, 84] | Matrix-based | Uses matrix preconditioning to improve optimisation efficiency. | Relevant when preconditioning is used to justify faster or more stable training. Report preconditioner cost, data scale, model scale, and compute-normalised robustness. |
| Ranger21<br>[85] | Hybrid | Combines multiple optimisation techniques into one training recipe. | Useful as a practical recipe but harder to attribute mechanistically. Report all enabled components, ablations where possible, tuning budget, and stress-test behaviour. |

##### Stress-test suite (minimal)

Evaluate both fidelity (e.g., PSNR/SSIM) and plausibility metrics under a small suite of domain-relevant corruptions (e.g., additive noise at multiple SNRs, blur/motion,intensity scaling/bias field [42, 61], changes in undersampling patterns when applicable, and small perturbations to conditioning inputs). Report sensitivity curves rather than single-point averages.

## Appendix C Expanded Tier Requirements and Claim Guardrails

### C.1 Tier definitions and required evidence

We propose a three-tier, compute-aware escalation to prevent overclaiming and ensure stability-first evidence proportional to deployment risk. Tier T1 is a low-overhead diagnostic tier intended for robustness characterization under irregular sampling and corruptions, but it is not sufficient for closed-loop or safety-critical deployment claims. Tier T2 adds moderate computation to stabilize optimization and support optimization-stability claims under stated shifts, by requiring curvature-aware training in at least one phase and reporting hazard-linked diagnostics such as *ρ*(*t*) and *T*_EoS_ aligned to the learning-rate schedule. Tier T3 is reserved for high-stakes settings and adds continuous-time inductive bias where irregular time is central (e.g., NODE/SSM or principled discretization) together with on-target deployment reporting (latency, memory, power proxy) and, when applicable, regime-shift evaluation such as lead time at fixed false alarm rate. Higher tiers subsume the requirements of lower tiers and justify correspondingly stronger claim scope.

#### Tier T1: Diagnose (minimal compute)

##### Claim scope

Evaluation-centric claims only: robustness characterization under irregular sampling and corruptions. Not eligible for safety-critical deployment or closed-loop claims.

##### Required checks

Irregular resampling/jitter; thinning and missingness; corruption suite appropriate to the modality (e.g., channel dropout and SNR sweeps for EEG; protocol/noise/motion for imaging); calibration (ECE) and selective prediction/abstention.

##### Must-report artifacts

Sensitivity curves (performance vs jitter/thinning/missingness); corruption curves; ECE; abstention rate vs input quality; at least one concrete failure analysis.

#### Tier T2: Stabilize optimization (moderate compute)

##### Claim scope

“Curvature-aware training improves robustness under stated shifts.” Limited deployment claims permitted only if runtime budgets are reported on target hardware.

##### Required checks

All T1 checks, plus curvature-aware optimization in at least one phase (e.g., AdamW pre-train then SAM/GCSAM fine-tune), and training stability diagnostics: *λ*_max_(*t*) (or defined surrogate), *ρ*(*t*) = *η*(*t*)*λ*_max_(*t*)*/*2, and *T*_EoS_ = Pr[*ρ*(*t*) *>* 1].

##### Must-report artifacts

Plots of *ρ*(*t*) and *T*_EoS_ aligned to *η*(*t*); metric *→* hazard*→* stress-test linkage statement; IID vs stress deltas reported side-by-side.

#### Tier T3: Continuous-time + deployment (high stakes)

##### Claim scope

Deployment-oriented stability claims under irregular observation and regime shifts, including edge/closed-loop, subject to stricter evidence.

##### Required checks

All T2 checks, plus continuous-time inductive bias where irregular time is central (e.g., Neural ODEs, SSMs, principled discretization), and deployment reporting (latency, memory, power proxy) on target hardware. For regime shift tasks, include the lead time at a fixed false alarm rate and the calibration of the warnings.

##### Must-report artifacts

Model-to-hardware mapping (solver/discretization details where relevant); on-target latency/memory/power; closed-loop safety gates (if applicable); stress diagnostics, including time perturbation and sensor-failure modes.

#### C.2 Claim-strength rule

To prevent over-claiming, papers must state the tier they satisfy and restrict conclusions accordingly: T1 supports robust evaluation claims; T2 supports optimization-stability claims; T3 supports deployment-oriented stability claims.

## Appendix D Theoretical Foundations of the Edge of Stability

### D.1 The Quadratic Approximation and Stability Threshold

The 2*/η* threshold arises from stability analysis of discrete-time descent in quadratic basins [11, 30, 62]. Near a local minimum *θ*^*⋆*^,

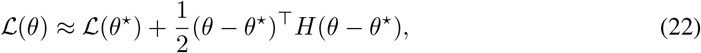

with gradient *∇ℒ* (*θ*) *≈ H*(*θ − θ*^*⋆*^) [11, 50]. The update implies

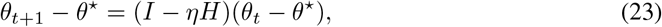

and strict stability requires |1 *− ηλ*_*i*_| *<* 1 for all eigenvalues *λ*_*i*_, hence *λ*_*i*_ *<* 2*/η* [49, 50].

*For completeness, θ*_*t*_ and *θ*_*t*_+_1_ denote the vectors of the model parameters in current and subsequent training iterations, *η* is the learning rate, *H* represents the Hessian matrix of the loss function, *I* denotes the identity matrix, and the superscript *⊤* indicates the vector transpose.

#### D.2 Qualitative phases of Edge-of-Stability training

Empirically, deep networks can exceed the 2*/η* boundary and still reduce loss, a phenomenon formalized as EoS [11, 30, 63]. Because computing the full Hessian is intractable, curvature is tracked via iterative [64, 65]. EoS training is often described in phases [11, 62, 65]:

1. **Progressive sharpening:** *λ*_max_ increases early in training.
2. **Crossing the threshold:** *λ*_max_ exceeds 2*/η*.
3. **EoS regime:** loss oscillates on short timescales while decreasing on longer timescales.
4. **Boundary-tracking regime:** *λ*_max_ remains near the stability boundary.

A common explanation is that globally non-convex loss landscapes allow unstable steps to leave local quadratic regions and enter flatter regions, producing a self-stabilizing trajectory [11, 64, 66, 67].

## Appendix E Supplementary Tables

**Table 6:**
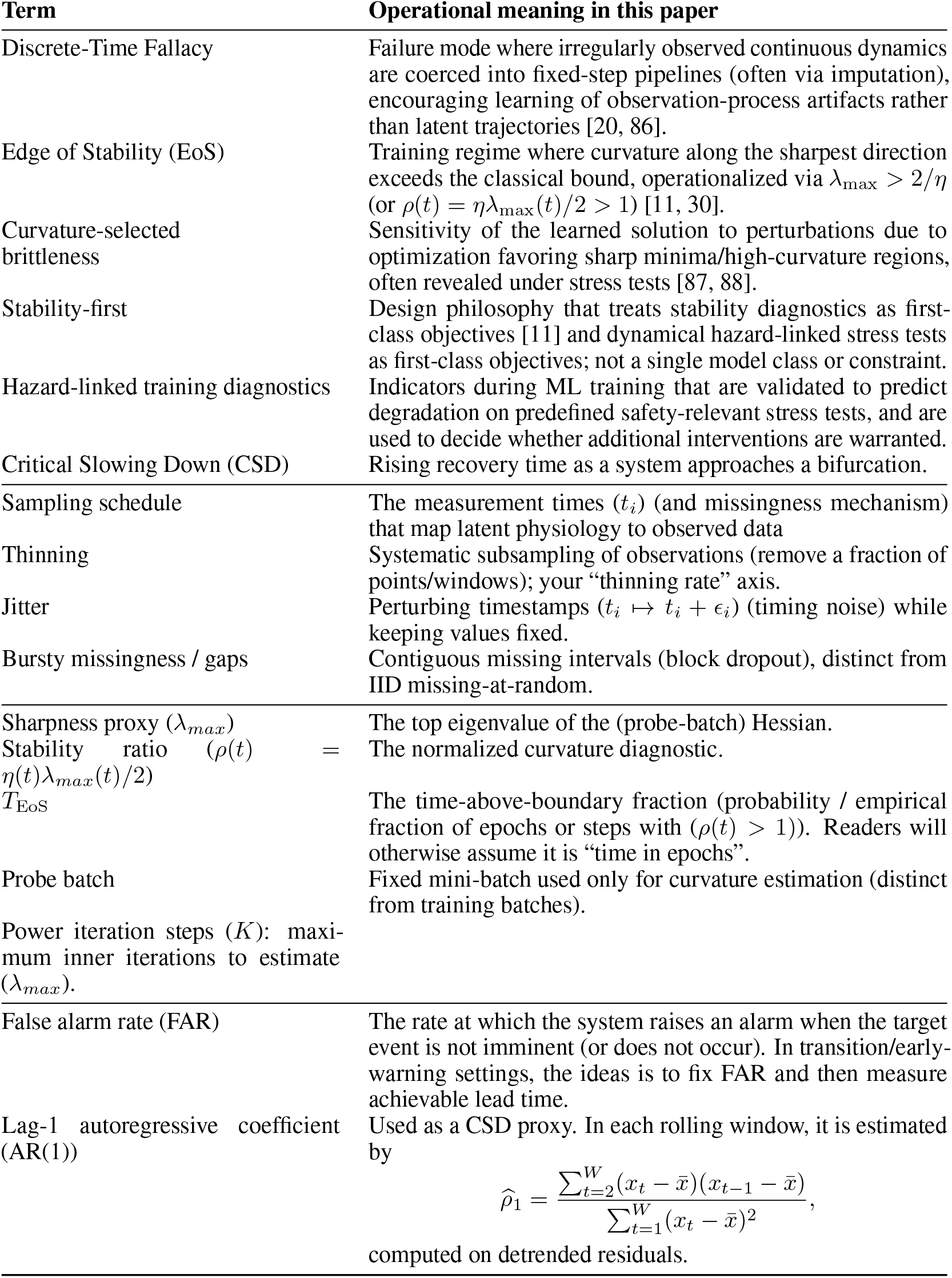
Glossary of terms and operational meanings.ss.

